# Post-COVID assessment in a specialist clinical service: a 12-month, single-centre analysis of symptoms and healthcare needs in 1325 individuals

**DOI:** 10.1101/2021.05.25.21257730

**Authors:** Melissa Heightman, Jai Prashar, Toby E Hillman, Michael Marks, Rebecca Livingston, Heidi Ridsdale, Kay Roy, Robert Bell, Michael Zandi, Patricia McNamara, Alisha Chauhan, Emma Denneny, Ronan Astin, Helen Purcell, Emily Attree, Lyth Hishmeh, Gordon Prescott, Rebecca Evans, Puja Mehta, Ewen Brennan, Jeremy Brown, Joanna Porter, Sarah Logan, Emma Wall, Hakim-Moulay Dehbi, Stephen Cone, Amitava Banerjee

**Affiliations:** University College London Hospitals NHS Trust, London, UK; University College London, London, UK; Clinical Research Department, Faculty of Infectious and Tropical Diseases, London School of Hygiene & Tropical Medicine, London, UK; Central and North West London NHS Foundation Trust, London, UK; Founder, UKDoctors#Longcovid; Member, Long COVID SOS; Lancashire Trials Unit, University of Central Lancashire, Preston; Francis Crick Institute, London, UK; Barts Health NHS Trust, London

## Abstract

**Background:** Complications following SARS-CoV-2 infection require simultaneous characterisation and management to plan policy and health system responses. We describe the 12-month experience of the first UK dedicated Post-COVID clinical service to include both hospitalised and non-hospitalised patients.

**Methods:** In a single-centre, observational analysis, we report outcomes for 1325 individuals assessed in the University College London Hospitals NHS Foundation Trust Post-COVID service between April 2020 and April 2021. Demography, symptoms, comorbidities, investigations, treatments, functional recovery, specialist referral and rehabilitation were compared by referral route (“post hospitalisation”, PH; “non-hospitalised”, NH; and “post emergency department”, PED). Symptoms associated with poor recovery or inability to return to work full-time were assessed using multivariable logistic regression.

**Findings:** 1325 individuals were assessed (PH 547 [41.3%], PED 212 [16%], NH 566 [42.7%]. Compared with PH and PED groups, NH were younger (median 44.6 [35.6-52.8] vs 58.3 [47.0-67.7] and 48.5 [39.4-55.7] years), more likely to be female (68.2%, 43.0% and 59.9%), less likely to be from an ethnic minority (30.9%, 52.7% and 41.0%) and seen later after symptom onset (median [IQR]:194 [118-298], 69 [51-111] and 76 [55-128] days) (all p<0.0001). NH patients had similar rates of onward specialist referral as PH and PED groups (18.7%, 16.1% and 18.9%, p=0.452), and were more likely to require support for breathlessness (23.7%, 5.5% and 15.1%, p<0.001) and fatigue (17.8%, 4.8%, 8.0%, p<0.001). Hospitalised patients had higher rates of pulmonary emboli, persistent lung interstitial abnormalities, and other organ impairment. 716 (54.0%) individuals reported <75% of optimal health (median [IQR] 70% [55%-85%]). Overall, less than half of employed individuals felt able to return to work full-time at first assessment.

**Interpretation:** Symptoms following SARS-CoV-2 infection were significant in both post- and non-hospitalised patients, with significant ongoing healthcare needs and utilisation. Trials of interventions and patient-centred pathways for diagnostic and treatment approaches are urgently required.

**Funding:** UCLH/UCL BRC

**Research in context:** *Previous evidence:* Long COVID and post-COVID syndrome were first identified in April 2020. We searched PubMed and medrxiv for articles published up to April 30th, 2021, using the keywords “long COVID”, “post-COVID syndrome”, “persistent symptoms”, “hospitalised”, “community” and “non-hospitalised”. We identified 17 articles and 7 systematic reviews. Fifteen studies have considered symptoms, multi-organ or functional impairment but only one study to-date has considered all these variables in non-hospitalised COVID patients. No studies have compared symptom burden and management between non-hospitalised and hospitalised individuals as systematically assessed and managed in a dedicated post-COVID service.

*Added value of this study:* For the first time, we report the baseline characteristics, investigation and outcomes of initial assessment of all eligible patients in a dedicated multi-professional post-COVID service, including 547 post-hospitalisation, 566 non-hospitalised and 212 patients discharged from emergency department. Despite relatively low comorbidity and risk factor burden in non-hospitalised patients, we show that both non-hospitalised and hospitalised patients presenting with persistent symptoms after SARS-CoV2 infection have high rates of functional impairment, specialist referral and rehabilitation, even 6-12 months after the acute infection. These real-world data will inform models of care during and beyond the pandemic.

*Implications of all the available evidence:* The significant, long-lasting health and social consequences of SARS-CoV-2 infection are not confined to those who required hospitalisation. As with other long-term conditions, care of patients experiencing Long COVID or specific end-organ effects require consistent, integrated, patient-centred approaches to investigation and management. At public health and policy level, burden of post-COVID morbidity demands renewed focus on effective infection suppression for all age groups.

## Introduction

Chronic post-viral sequelae are well-known(1). The SARS-CoV-2 pandemic’s scale therefore poses an unprecedented threat to long-term health(2,3). Initially, funding, research, clinical practice and policy emphasised acute, hospitalised patients in areas with a high burden of COVID-19 cases and deaths. However, more recently, there has been increasing focus on longer-term effects of acute infection (“Long COVID” or “post-COVID syndrome”), including non-hospitalised (NH) patients(4, 5) (**Figure 1**).

**Figure 1.**
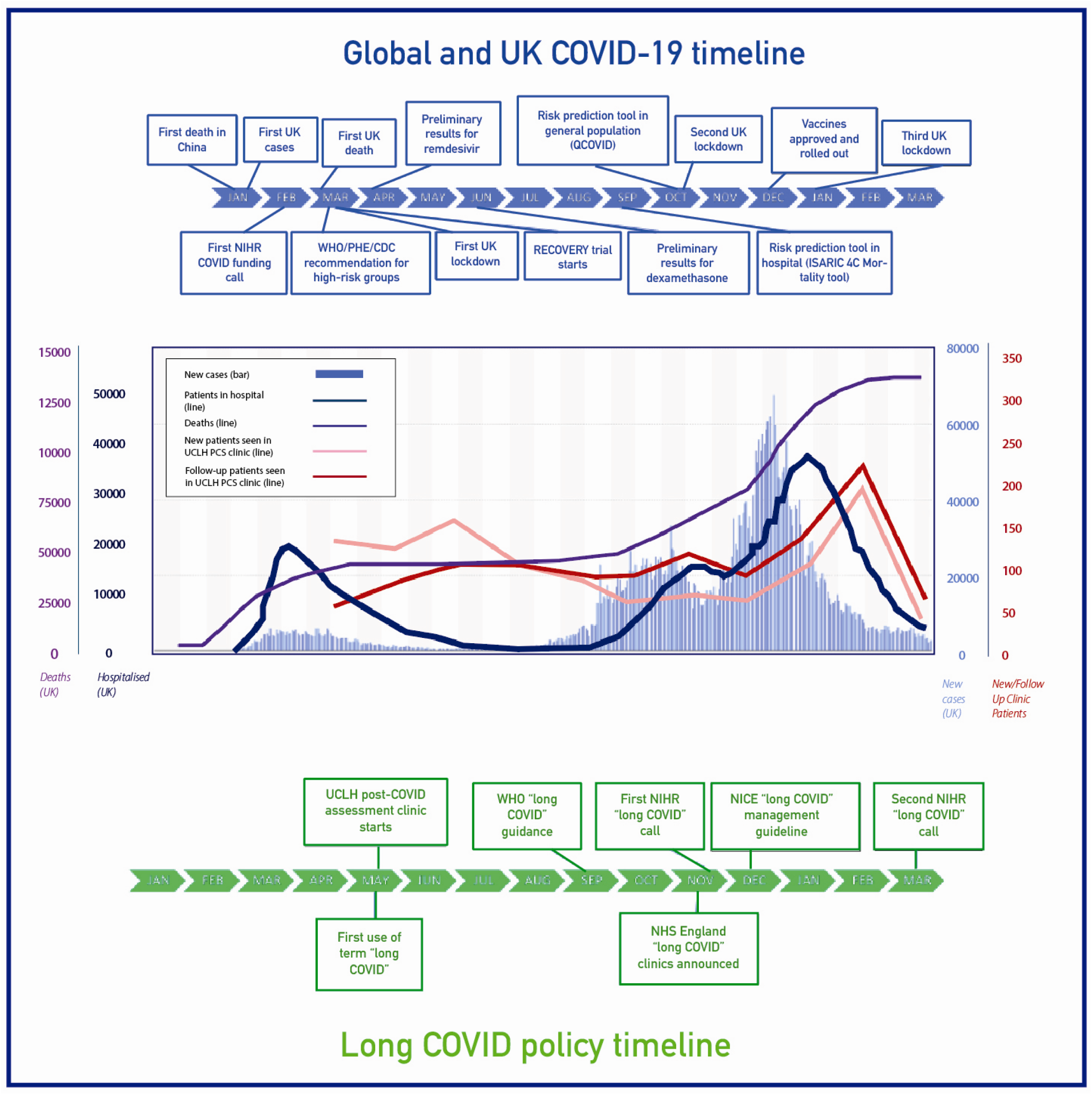
Post-COVID assessment in the context of the pandemic.

Clinically, Long COVID remains a poorly defined disease (5,6). To address the healthcare needs of the estimated 1.1 million individuals with Long COVID in the UK(7) and the millions around the world(8) there is a need to deliver clinical care whilst simultaneously reviewing clinical data in “learning health system” approaches(9). Research on clinical characterisation and management has been identified by both recent expert consensus groups and patients as a priority area(10).

COVID-19 hospitalisation is associated with significant risk of end-organ impact, functional impairment, readmission, and mortality(11-13). Recent analysis of 6-month post-COVID outcomes in China highlighted persistent physical, mental and functional impact during follow-up but excluded non-hospitalised (NH) patients(14). Recent electronic health record (EHR) analyses in the US and Denmark respectively suggested major healthcare resource implications of Long COVID in NH individuals(15,16), but no studies to-date have reported on models of care for Long COVID.

More than 80 dedicated Post-COVID assessment clinics have been announced in England(17), but many centres only started accepting referrals one year after the first pandemic wave. Various care models have been proposed through expert and patient consensus(18-19), but real world data are lacking, particularly in community settings, where the majority of COVID-19 and Long COVID patients are managed.

In April 2020, we established a dedicated service for assessment of post-COVID complications at University College London Hospitals NHS Foundation Trust(UCLH) for both hospitalised and non-hospitalised individuals. We report baseline characteristics, clinical presentation, management and outcomes for all individuals referred to this specialist clinic following suspected or confirmed SARS-CoV-2 infection over a 12-month period.

## Methods

### Context of the UCLH post-COVID service

The UCLH post-COVID service is a one-stop model of assessment (by physician and physiotherapist), diagnostics and exercise test to triage need for further specialist input, treatment or rehabilitation. It accepts referrals from three sources: (i)*Post-hospitalised(PH)*: post-admission to UCLH with COVID-19; (ii)*Non-hospitalised(NH)*: individuals referred from primary care with suspected long COVID ≥6 weeks post-SARS-CoV-2 infection, and (iii)*Post*-*emergency department(PED)* referral for individuals with persistent symptoms at 4-6 weeks after attendance. As per British Thoracic Society Guidance(20) post-discharge review in PH patients was at 6 weeks for those who received respiratory support via continuous positive airway pressure (CPAP) or invasive ventilation, or with chest imaging abnormality, and at 12 weeks in all other patients. PH patients under care of other specialist services or without chest X-ray abnormalities were not routinely booked into the post-COVID service.

### Clinic population

Our analysis included all patients assessed in the UCLH post-COVID service between April 20, 2020 and April 25, 2021**(Figure 2)**, excluding follow-up assessments and individuals who did not attend. Due to restricted access to testing during the first pandemic wave, SARS-CoV-2 infection was defined by either laboratory confirmation (viral positive oropharyngeal/nasopharyngeal swab when tested by reverse-transcriptase PCR, or anti-N antigen IgG detected in convalescent serum), or strong clinical suspicion (assessed by both referring and consulting clinicians)(21).

**Figure 2.**
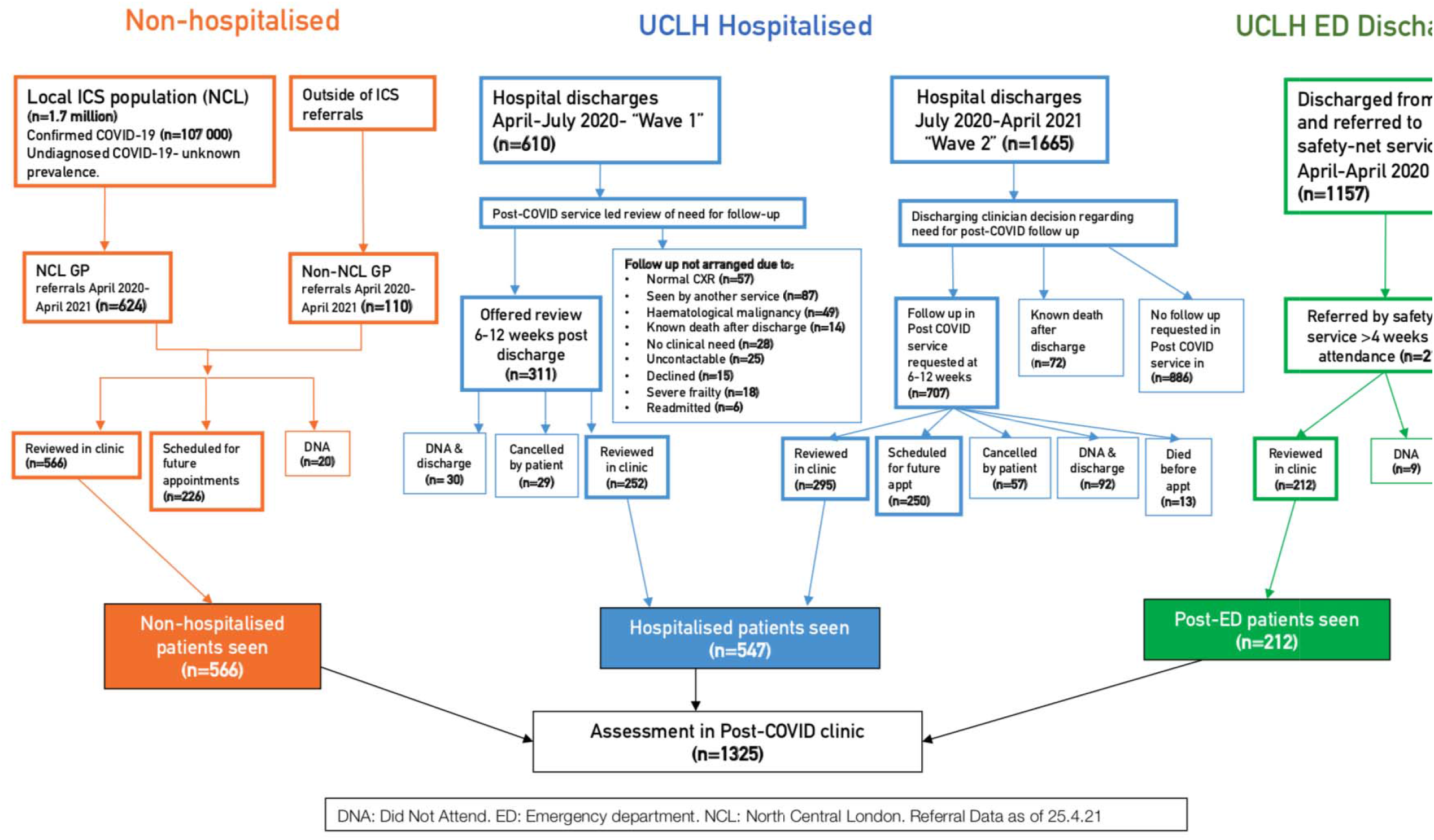
Study Population undergoing assessment in post-COVID assessment clinic.

### Post-COVID Assessment

Clinical assessment was developed by secondary care clinicians and therapists, consisting of a consultation and multi-professional assessment, delivered primarily face-to-face, or where necessary, virtually. An EHR (Epic™- Epic Systems Corporation, Wisconsin) structured assessment tool (accessible via patient portal before appointments and during assessment) was used to record socio-demography, past medical history, current symptoms and functional status. Where appropriate, the following outcome measures were recorded: percentage of best health (as used in other tools e.g. EuroQuol-5 domain-5 level), symptom severity for breathlessness, fatigue, cough, sleep disturbance and palpitations, MRC Dyspnoea scale, post-traumatic stress disorder scale(PTSD), Fatigue Assessment Scale(FAS), Generalised Anxiety Disorder 2-item(GAD2) and Patient Health Questionnaire 2-item(PHQ2)(5,11,13,21) (**Web Methods Supplement**).

Selected patients underwent further investigation at discretion of the clinician or following multi-disciplinary team meetings with respiratory, cardiology and neurology input according to clinical need. These tests included: full blood count, liver and renal function, D-dimer, troponin and NT-pro-brain-natriuretic peptide(NT-proBNP); sit-to-stand testing; chest X-ray; CT pulmonary angiography with HRCT pre-contrast(CTPA); electrocardiogram(ECG); cardiac magnetic resonance imaging(cMRI); brain MRI; echocardiography; and Holter monitoring. Lung function testing was unavailable due to local infection control requirements.

### Clinical Management

After assessment, patients were either discharged to the community, booked for further clinic follow-up, and/or referred for specialist opinion, physical rehabilitation, respiratory physiotherapy, fatigue management, vocational support and psychology support. Patients with elevated GAD or PHQ scores (≥3) were advised on self-referral to “improving access to psychological therapies” (iAPT) services.

### Data extraction and statistical analysis

Demographic and clinical data at first assessment were extracted from the EHR. Age, time since symptom onset in days, self-reported percentage of best health were recorded as continuous variables, and the presence or absence of individual symptoms as binary variables. Self-reported ability to return to work, for employed individuals, was recorded on an ordinal scale (‘Not at all’ to ‘Full time’). The Index of Multiple Deprivation(IMD) decile was derived from each patients’ post-code. We use descriptive statistics to summarise baseline characteristics. Continuous variables are reported as median with interquartile ranges (IQR), categorical variables are reported as frequency (%). For group-wise comparison, we use the Kruskal-Wallis test for continuous variables and the chi-squared test or Fisher’s exact test for categorical variables. We used multivariable logistic regression models, adjusted for age (modelled as a continuous variable) and gender (male vs. female as reference group), to investigate symptoms associated with a) optimal (≥75%) patient-reported functional recovery at first presentation and b) patient-reported ability to return to full-time employment at first presentation. The model for returning to employment excluded those not employed or retired before Covid. Age and gender were included in the models and all recorded symptoms (represented as presence vs. absence) were available for selection in a backwards stepwise selection process with a threshold of p<0.05. A sensitivity analysis also considered time since onset for selection in the models. Referral rates of local GP practices (per 1000 practice population) were determined using locally available EHR data for practice size and referring practice. All analyses were performed using Python 3.7.6.

### Ethics

This study was confirmed to be a service evaluation, and ethical approval was waived, by UCLH’s Research and Development and Information Governance Directorates as part of the UCLH Data Access Committee set up in response to the COVID-19 Pandemic (https://www.uclhospitals.brc.nihr.ac.uk/clinical-research-informatics-unit/data-explorer), following UK COVID-19 guidelines for use of patient data (https://www.hra.nhs.uk/covid-19-research/guidance-using-patient-data/).

## Results

### Socio-demographic profile

The number of referrals to the UCLH Post-COVID clinic mirrored successive pandemic waves(**Figure 1**). Excluding patients who did not attend and cancelled appointments, 1325 patients were reviewed: PH(n=547), NH(n=566) and PED(n=212)(**Figure 2**). 614(46.3%) patients were tested for SARS-CoV-2 infection using RT-PCR, of which 378(61.6%) were positive, and serological testing (n=241;18.2%) with 114(47.3%) positive. The remaining 470(35.5%) patients had strong clinical suspicion of prior SARS-CoV-2 infection.

Median age was 49.9 [IQR 40.1-60.1] years, 748 (56.5%) patients were women and 550 (41.5%) were of non-white ethnicity. Compared with PH and PED individuals, NH were younger (44.6 [IQR 35.6-52.8] years vs 58.3 [IQR 47.0-67.7] and 48.5 [IQR 39.4-55.7], p<0.001), more likely to be female (68.2% vs 43.0% and 59.9%, p<0.001), less likely to be non-white (30.9% vs 52.7% and 41.0%, p<0.001) and less likely to live in an area of social deprivation (IMD category, median [IQR]: 5[3-7] vs 4[2-6] and 4[3-6], p<0.001) (**Table 1, Web Figure 1**). General practice referral rates in the catchment population ranged from 0.09-3.64 patients per 1000 practice population, with 49/201 (24.4%) of practices referring no patients (**Web figure 2, Supplementary file**).

**Table 1:**
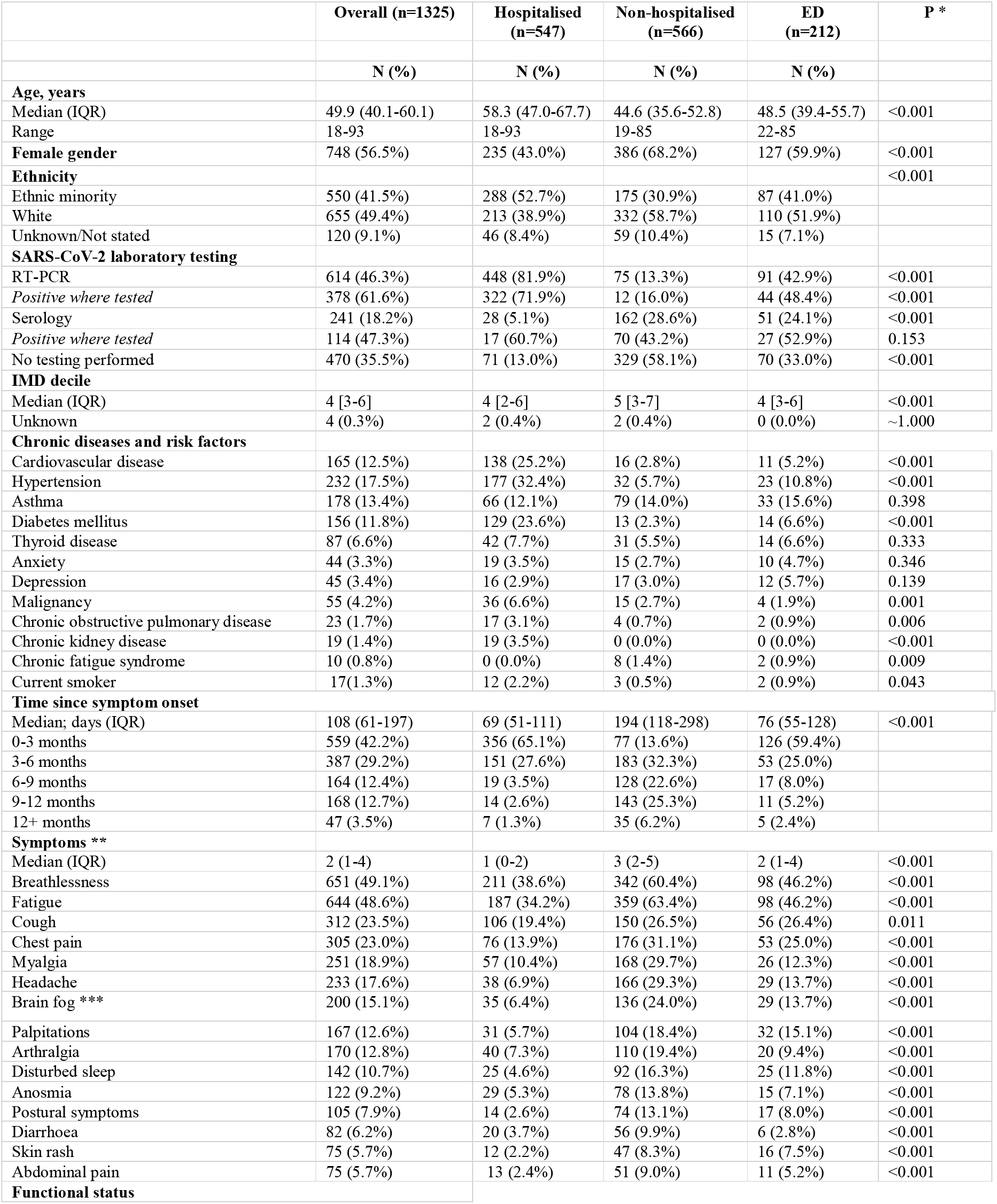
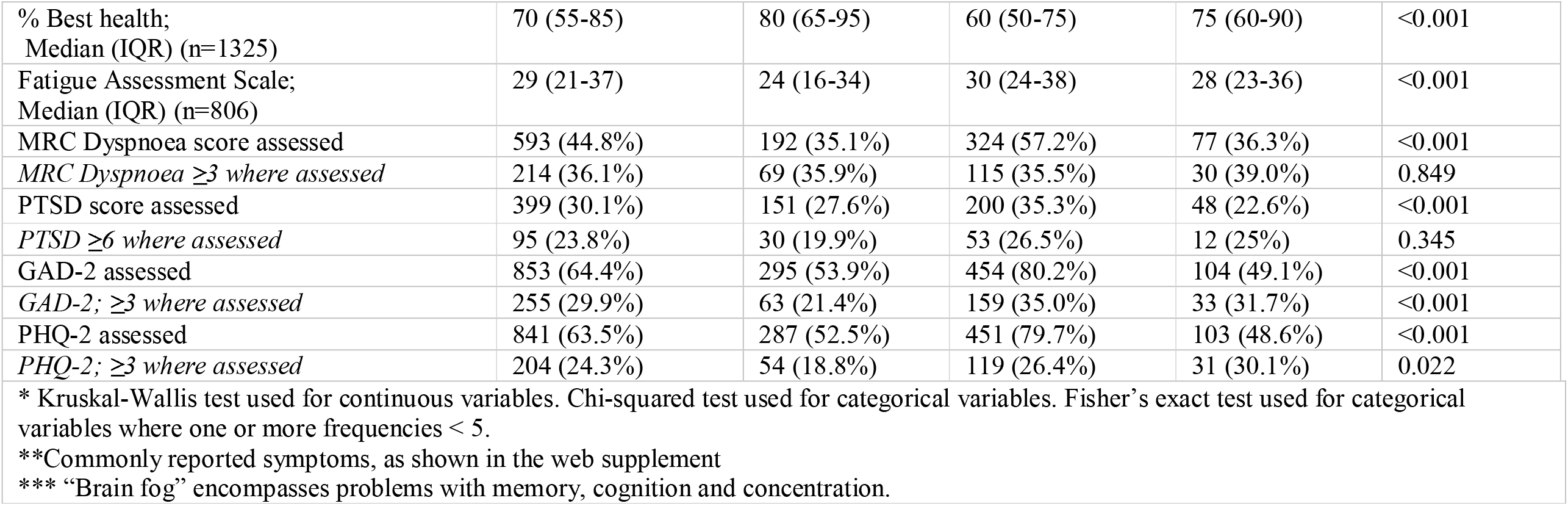
Baseline characteristics in 1325 individuals referred to the post-COVID assessment clinic.

### Chronic diseases and risk factors

Hypertension (17.5%), asthma (13.4%), cardiovascular disease (12.5%), diabetes (11.8%) and thyroid disease (6.6%) were the most common comorbidities. Most pre-morbid chronic diseases were more common amongst PH patients compared with NH and PED patients. The exceptions were asthma (12.1% vs 14.0% and 15.6%), anxiety (3.5% vs 2.7% and 4.7%), depression (2.9% vs 3.0% and 5.7%) and chronic fatigue syndrome (0.0% vs 1.4% and 0.9%). Current smoking was uncommon (1.3% overall) (**Table 1**). Overall, 86 PH patients died before a post-COVID clinic assessment, with a further two PH patients dying after clinic review.

### Symptoms and functional status

Time from symptom onset (of acute SARS-COV-2 infection) to first clinic assessment was a median of 108 days [IQR 61-197], and delayed in NH (194 days [118-298] compared with PH (69[51-111] days) and PED patients (76 [55-128] days) (p<0.001).

At first visit, median [IQR] number of reported symptoms was 2[1-4] overall (PH, NH and PED groups: 1[0-2], 3[2-5] and 2[1-4], p<0.001). Most commonly reported symptoms were breathlessness (49.1%), fatigue (48.6%), cough (23.5%), myalgia (18.9%), chest pain (23.0%), headache (17.6%), “brain fog” (15.1%) and palpitations (12.6%). All symptoms were more frequently reported by NH than PH and PED patients (**Table 1**). Overall, 36.1% had MRC dyspnoea scale scores ≥3 (n=593 assessed), 23.8% PTSD score >6 (n=399 assessed), 29.9% GAD-2 ≥3 (n=853 assessed), and 24.3% PHQ2 ≥3 (n=841 assessed), with significant differences (p<0.001) between referral sources (**Table 1**). Breathlessness, fatigue, palpitations, sleep quality and chest pain were more common and, where reported, rated as more severe (**Web Table 3**) in NH compared to PH groups. Symptom co-occurrence varied between groups **(Figure 3)**. At first assessment, 716 (54.0%) of individuals reported <75% of optimal health, more frequently in NH (71.8%) compared with PED (37.8%) and PH (48.6%) individuals **(Web Table 1)**.

**Figure 3.**
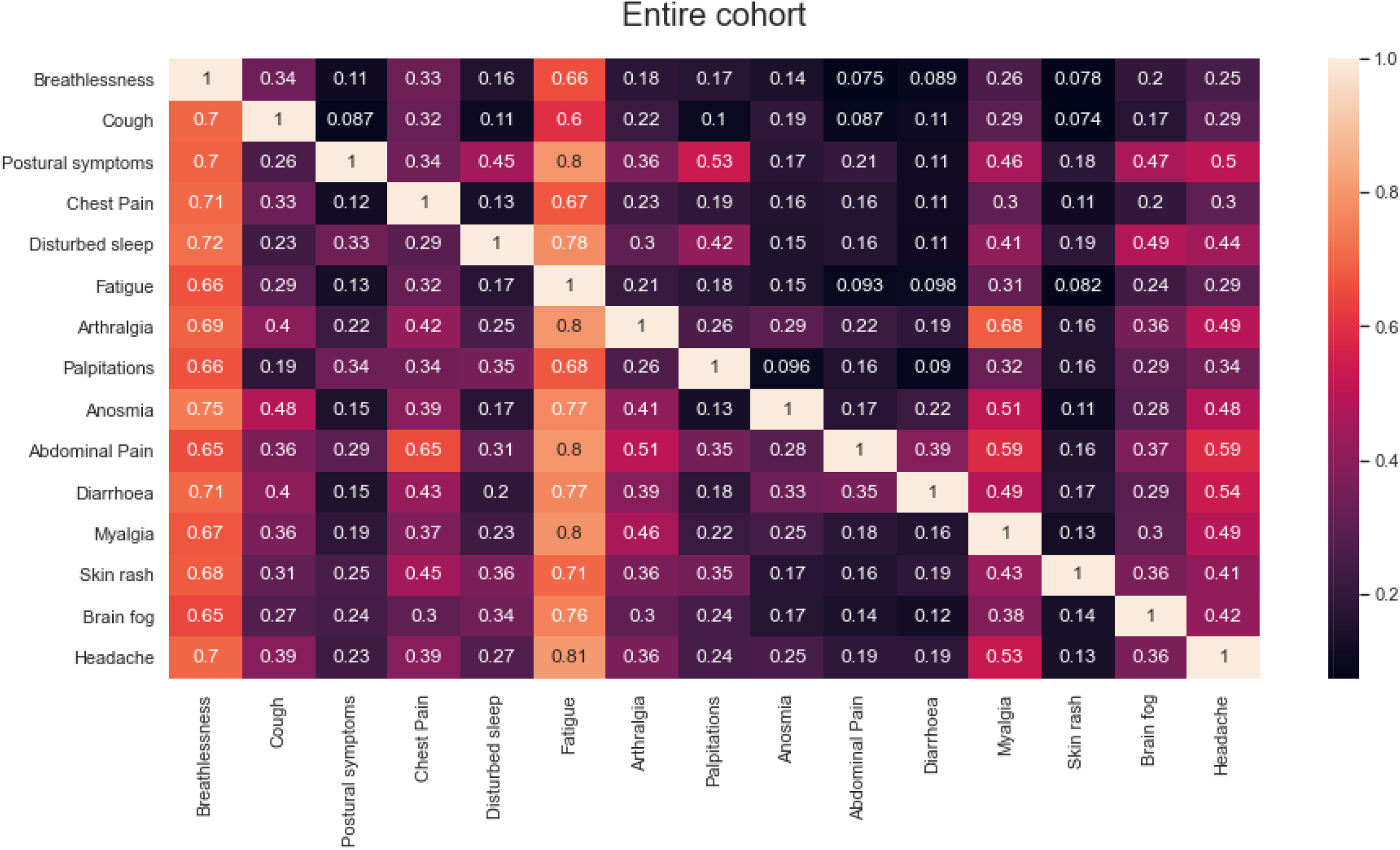
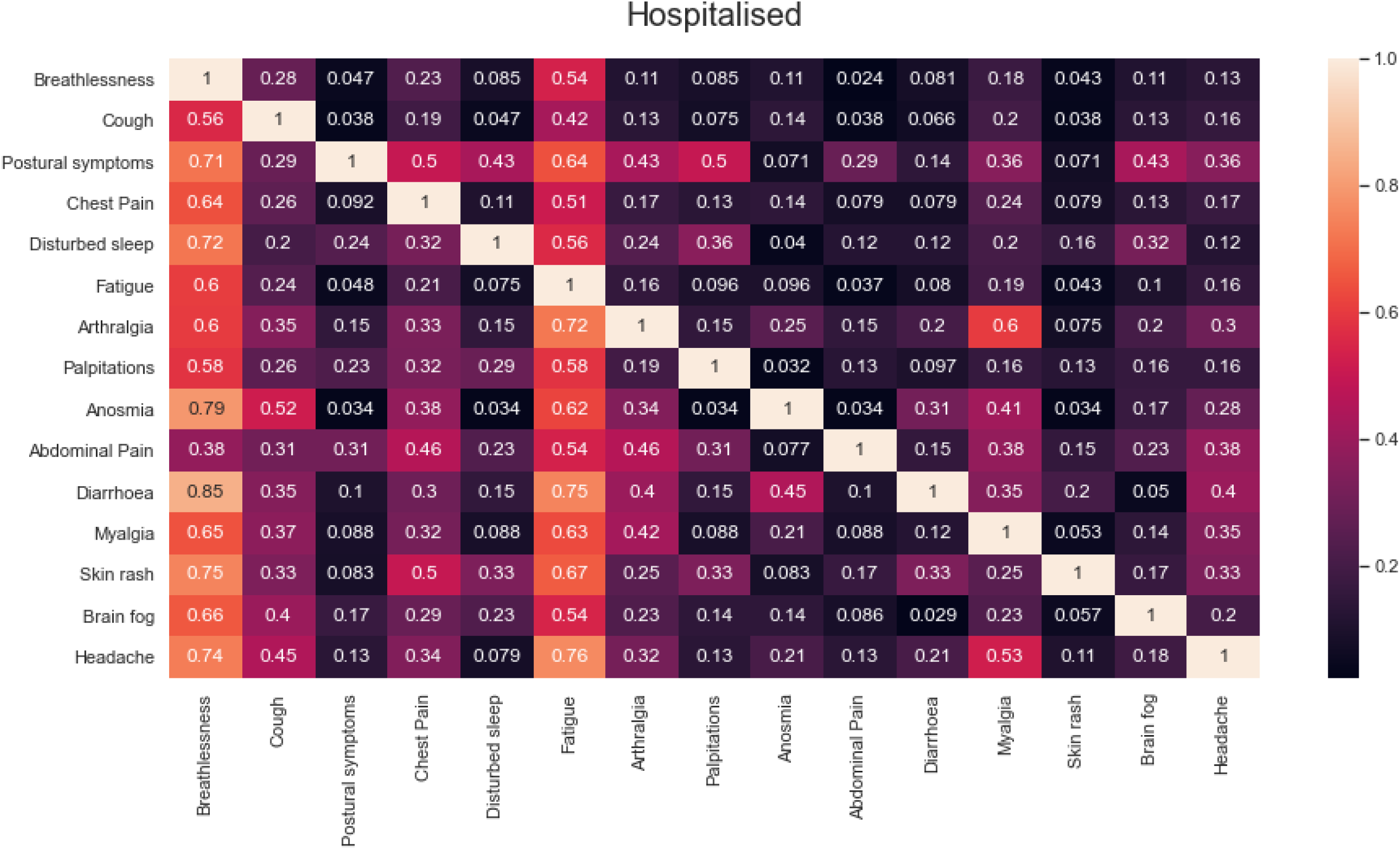
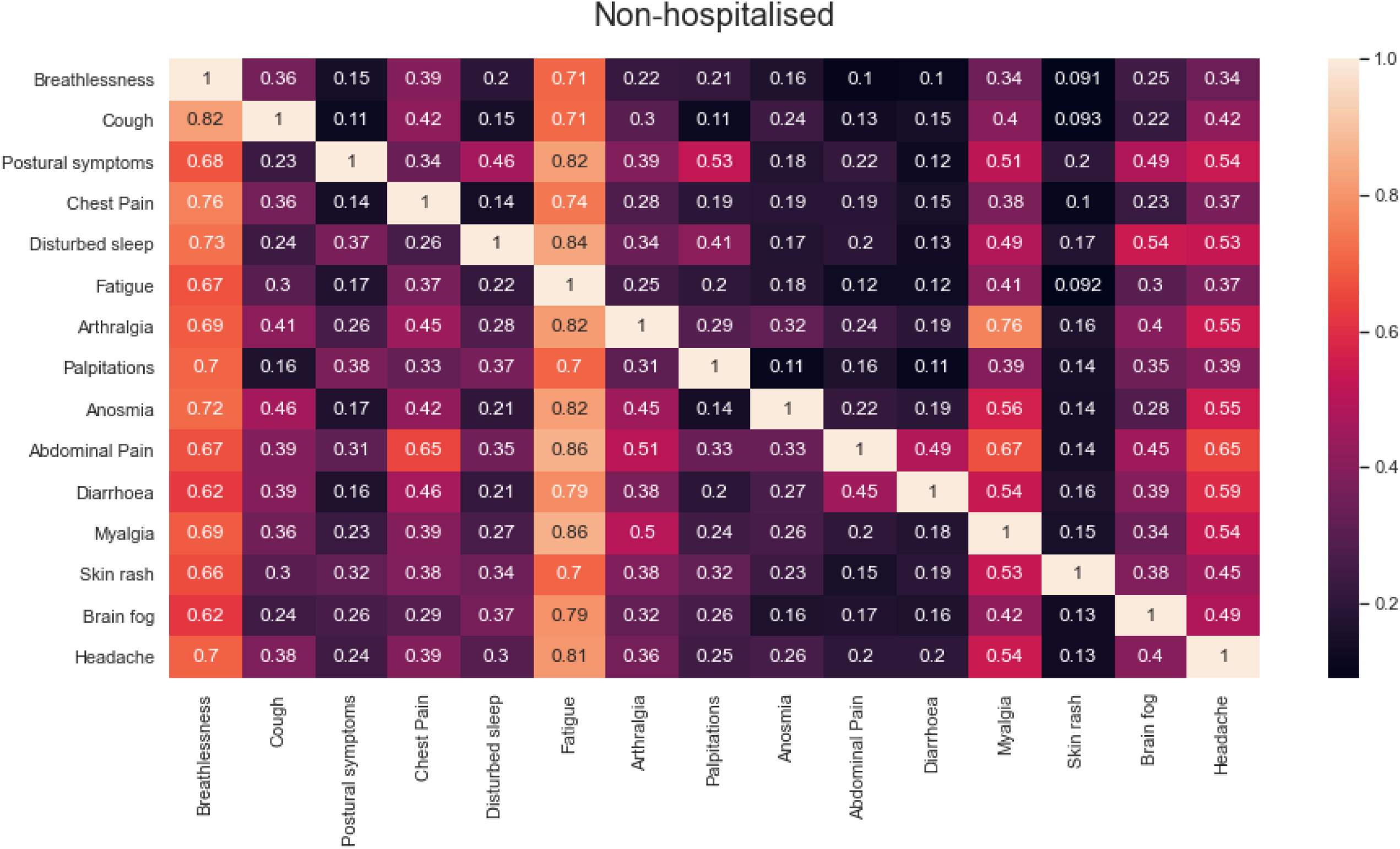
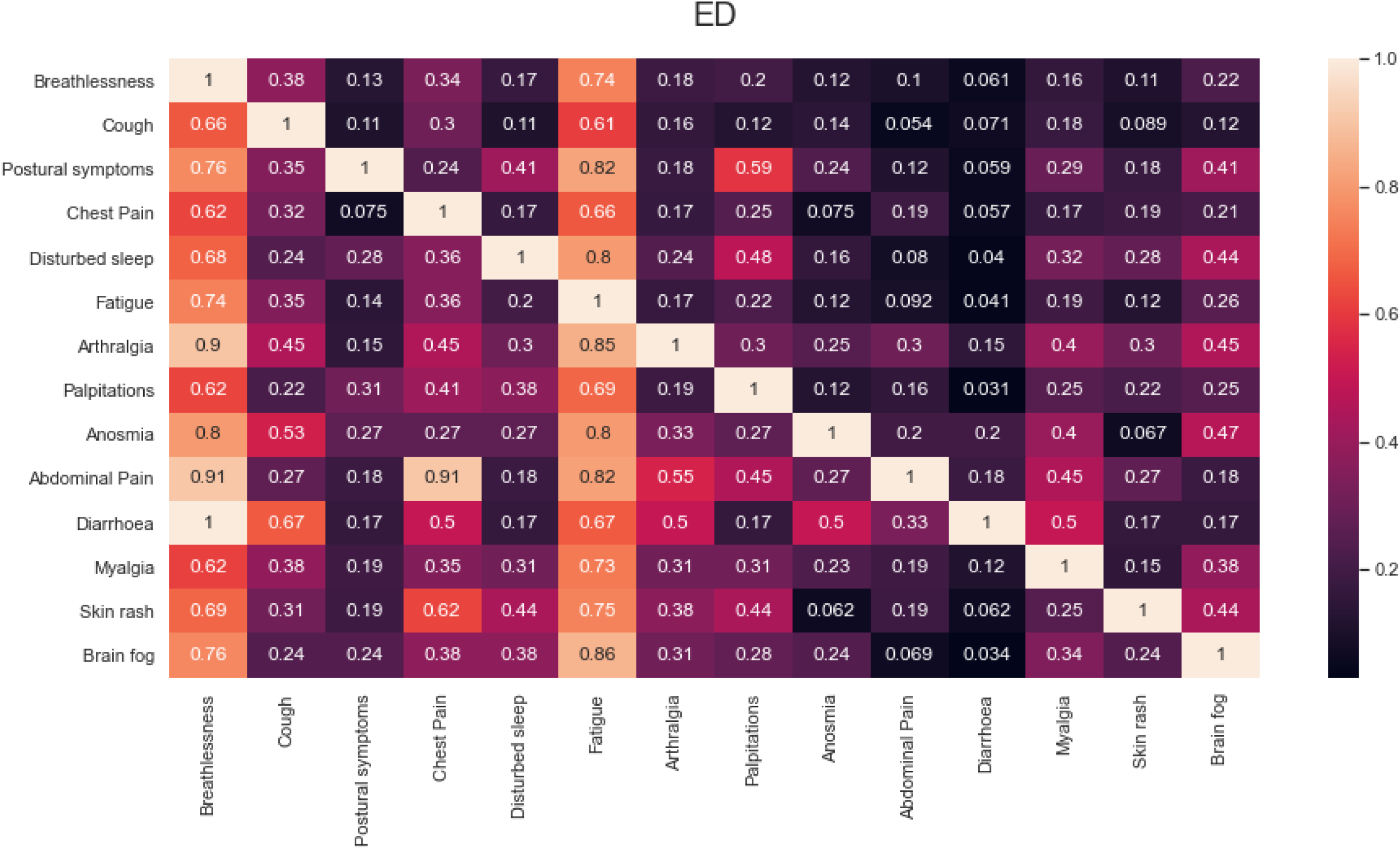
Co-occurrence of symptoms at first assessment in 1325 individuals referred to the post-COVID assessment clinic.

### Investigations

Investigations requested according to clinician judgement included chest X-ray(n=694, 52.4%), echocardiography(n=330, 24.9%), Holter monitor (n=222, 16.8%), CTPA(n=204, 15.4%), 6-minute walk tests(n=142, 10.7%), cMRI (for chest pain or troponin elevation, n=76, 5.7%) and MRI brain imaging(n=40, 3.0%) (**Table 2**). Of 204 CTPAs requested, 5.9%(0.9% of cohort) showed PE and 30.9%(4.8% of cohort) showed persistent lung interstitial changes. Outside the post-COVID service, 8 patients had PE identified via acute medical services; and 49 (9.0%) PH patients had PE on CTPA during inpatient stay. Echocardiography showed left ventricular systolic dysfunction in 7 (2.2% of those examined). cMRI showed mild myocarditis in 24(31.6% of those scanned) and evidence of ischaemic heart disease in 11 PH patients(35.5% of those scanned). On MRI brain, no changes attributed to COVID were identified. Where tested, NT-proBNP was raised in 8.0% of individuals, absolute eosinophil count in 5.8%, troponin-T in 13.4%, creatine kinase in 16.9%, alanine transaminase in 18.0% and D-dimer in 18.9%. Abnormal blood investigations were more common in PH than NH individuals(D-dimer: 28.3% vs 10.3%; troponin T: 27.0% vs 4.7%; NT-proBNP: 17.0% vs 0.0%; and eosinophil count: 6.8% vs 2.5%)(**Web Table 2**).

**Table 2.** Investigations, outcomes and onward referrals in 1325 individuals referred to the post-COVID assessment clinic.

|  | Overall<br>(n=1325) | Hospitalised<br>(n=547) | Non-hospitalised<br>(n=566) | ED<br>(n=212) | P* |
| --- | --- | --- | --- | --- | --- |
|  | N (%) | N (%) | N (%) | N (%) |  |
| **Investigations** |  |  |  |  |  |
| Chest X-ray | 694 (52.4%) | 355 (64.9%) | 204 (36.0%) | 135 (63.7%) | <0.001 |
| CT pulmonary angiogram with HRCT pre-contrast (CTPA) | 204 (15.4%) | 104 (19.0%) | 64 (11.3%) | 36 (17.0%) | 0.001 |
| *Pulmonary embolism (PE) detected in scans performed* | 12 (5.9%) | 9 (8.7%) | 1 (1.6%) | 2 (5.6%) | 0.159 |
| *Persistent interstitial abnormalities in scans performed* | 63 (30.9%) | 49 (47.1%) | 8 (12.5%) | 6 (16.7%) | <0.001 |
| Echocardiogram | 330 (24.9%) | 121 (22.1%) | 160 (28.3%) | 49 (23.1%) | 0.048 |
| *Ejection Fraction <55% in investigations performed.* | 7 (2.1%) | 3 (2.5%) | 4 (2.5%) | 0 (0.0%) | 0.763 |
| Holter | 222 (16.8%) | 51 (9.3%) | 143 (25.3%) | 28 (13.2%) | <0.001 |
| 6-minute walk test | 142 (10.7%) | 24 (4.4%) | 98 (17.3%) | 20 (9.4%) | <0.001 |
| Cardiac MRI | 76 (5.7%) | 31 (5.7%) | 33 (5.8%) | 12 (5.7%) | 0.992 |
| *Mild myocarditis in scans performed* | 24 (31.6%) | 9 (29.0%) | 15 (45.5%) | 0 (0%) | 0.008 |
| MRI Brain | 40 (3.0%) | 27 (4.9%) | 9 (1.6%) | 4 (1.9%) | 0.004 |
| Lung function | 17 (1.3%) | 8 (1.5%) | 9 (1.6%) | 0 (0.0%) | 0.171 |
| Sleep study | 8 (0.6%) | 3 (0.5%) | 4 (0.7%) | 1 (0.5%) | ~1.000 |
| Sit-to-stand test | 749 (56.5%) | 238 (43.5%) | 415 (73.3%) | 96 (45.3%) | <0.001 |
| *Post-test oxygen saturation ≤92% in tests undertaken.* | 58 (7.7%) | 35 (14.7%) | 18 (4.3%) | 5 (5.2%) | <0.001 |
| Discharged at first appointment | 98 (58-170) | 63 (46-101) | 183 (118-329) | 72 (51-112) | <0.001 |
| At least one follow-up visit | 126 (70-221) | 80 (59-126) | 198 (124-274) | 89 (58-172) | <0.001 |
| **First visit outcomes** |  |  |  |  |  |
| Discharged from clinic | 740 (55.8%) | 338 (61.8%) | 288 (50.9%) | 114 (53.8%) | <0.001 |
| Specialist referral | 234 (17.7%) | 88 (16.1%) | 106 (18.7%) | 40 (18.9%) | 0.452 |
| Seen by physiotherapist | 776 (58.6%) | 236 (43.1%) | 398 (70.3%) | 142 (67.0%) | <0.001 |
| Psychology referrals to iAPT | *iAPT- improving adult access to psychological therapies.* Advised if GAD or PHQ elevated (see Table 1) – patients can self refer to this service |  |  |  |  |
| **Specialist referral** |  |  |  |  |  |
| Cardiology | 110 (8.3%) | 41 (7.5%) | 46 (8.1%) | 23 (10.8%) | 0.317 |
| Neurology | 125 (9.4%) | 26 (4.8%) | 87 (15.4%) | 12 (5.7%) | <0.001 |
| ENT | 31 (2.3%) | 4 (0.7%) | 20 (3.5%) | 7 (3.3%) | 0.002 |
| Other | 53 (4.0%) | 19 (3.5%) | 24 (4.2%) | 10 (4.7%) | 0.683 |
| **Physiotherapist outcomes** | **n=776**<br>**(58.6%)** | **n=236 (43.1%)** | **n=398**<br>**(70.3%)** | **n=142**<br>**(67.0%)** |  |
| Discharge with self-management support ** | 363 (46.8%) | 133 (56.4%) | 152 (38.2%) | 78 (54.9%) | <0.001 |
| Cognitive rehabilitation ** | 17 (2.2%) | 5 (2.1%) | 9 (2.3%) | 3 (2.1%) | ~1.000 |
| Speech and language therapy ** | 46 (5.9%) | 31 (13.1%) | 7 (1.8%) | 8 (5.6%) | <0.001 |
| Respiratory physiotherapy** (including ENO***) | 196 (25.3%) | 30 (12.7%) | 134 (33.7%) | 32 (22.5%) | <0.001 |
| Fatigue management ** | 144 (18.6%) | 26 (11.0%) | 101 (25.4%) | 17 (12.0%) | <0.001 |
| * Kruskal-Wallis test for continuous variables. Chi-squared test for categorical variables. Fisher's exact test used for categorical variables where one or more frequencies < 5.<br>** Percentages reported as % of individuals seen by physiotherapist.<br>*** ENO- English National Opera "Breathe" program. |  |  |  |  |  |

### Outcomes and onward referrals

740(55.8%) individuals were discharged after first assessment(61.8%, 50.9% and 53.8% in PH, NH, and PED). Individuals who required follow-up had longer symptom duration at presentation (median 126, IQR 70-221 days) than those discharged at first visit (median 98[58-170] days). PH, NH and PED individuals had similar specialist referral rates (16.1%, 18.7% and 18.9%, p=0.452), most commonly to cardiology(8.3%) and neurology(9.4%)(**Table 2**). Of 776(58.6% of study population) individuals assessed by physiotherapists, 363(46.8%) were discharged with self-management support at first visit. NH were more likely than PH and PED individuals to require support for disordered breathing pattern(23.7%, 5.5% and 15.1%, p<0.001) and referral for fatigue management (25.4%, 11.0% and 12.0%, p<0.001) (**Table 2**).

### Optimal health and employment

In PH, NH, and PED groups, the median self-reported proportion with optimal health was 80% [65-95%], 60% [50-75%] and 75% [60-90%] respectively, correlating negatively with symptom duration at time of assessment (**Table 1 and Web Figure 3**). Overall, less than half of employed individuals felt able to return to work full-time at first assessment (**Web Table 1**).

In multivariable logistic regression analysis(**Table 3**), younger age was associated with return to work(PH, NH) and male gender(PH, ED). For PH individuals, fatigue, brain fog, chest pain and breathlessness were associated with inability to work full-time. Arthralgia and headache were associated with full-time return to work. For NH individuals, fatigue, brain fog, and headache were associated with inability to return to work. For the PED group, breathlessness and myalgia were associated with non-return to work, whereas cough and arthralgia were associated with return to work. For some included symptoms, 95% confidence intervals are wide, suggesting uncertainty about their effect, perhaps due to small numbers of patients with that symptom. Older age and male gender were associated with return to optimal health status (≥75%). Fatigue and postural symptoms were significantly associated with suboptimal (<75%) health status in PH and NH groups, and brain fog in NH and PED groups(**Table 3**). Postural symptoms were rare in PH patients, and few with this symptom achieved optimal health. The extreme odds ratio probably reflected the small number recovering in this group, rather than importance of this symptom. Sensitivity analyses including time since onset in the backwards selection process only changed models for recovery in NH patients, where longer times since onset and disturbed sleep were associated with lower odds of recovery, replacing postural symptoms in the model.

**Table 3.** Multivariable logistic regression analysis showing symptoms associated with ability to return to work full-time and ≥75% functional recovery at first assessment in 1325 individuals referred to the post-COVID assessment clinic.

| | Return to work full-time<br>(n=1028) | | | $\geq 75\%$ functional recovery<br>(n=1325) | | |
| --- | --- | --- | --- | --- | --- | --- |
|  | Covariate | Multivariable OR<br>(95% CI) | P | Covariate | Multivariable<br>OR (95% CI) | P |
| **Hospitalised** |  |  |  |  |  |  |
|  | Age | 0.97 (0.95-0.99) | 0.008 | Age | 1.01 (1.00-1.03) | 0.019 |
|  | Male gender | 1.88 (1.33-2.67) | <0.001 | Male gender | 2.58 (1.94-3.42) | <0.001 |
|  | Brain fog | 0.13 (0.03-0.58) | 0.008 | Postural<br>symptoms | 0.05 (0.01-0.46) | 0.007 |
|  | Chest pain | 0.28 (0.13-0.60) | 0.001 | Chest pain | 0.43 (0.25-0.74) | 0.002 |
|  | Fatigue | 0.29 (0.17-0.52) | <0.001 | Fatigue | 0.47 (0.33-0.68) | <0.001 |
|  | Breathlessness | 0.54 (0.33-0.90) | 0.019 | Arthralgia | 2.69 (1.22-5.92) | 0.014 |
|  | Arthralgia | 2.55 (1.01-6.42) | 0.048 |  |  |  |
|  | Headache | 2.75 (1.04-7.25) | 0.041 |  |  |  |
| **Non-hospitalised** |  |  |  |  |  |  |
|  | Age | 0.98 (0.96-0.99) | 0.008 | Age | 1.02 (1.00-1.04) | 0.018 |
|  | Male gender | 1.20 (0.84-1.74) | 0.319 | Male gender | 1.44 (0.99-2.09) | 0.058 |
|  | Brain fog | 0.54 (0.35-0.86) | 0.008 | Postural<br>symptoms | 0.08 (0.02-0.32) | <0.001 |
|  | Headache | 0.64 (0.42-0.97) | 0.034 | Fatigue | 0.49 (0.35-0.68) | <0.001 |
|  | Fatigue | 0.67 (0.5-0.92) | 0.012 | Myalgia | 0.49 (0.30-0.81) | 0.005 |
|  |  |  |  | Brain fog | 0.53 (0.31-0.89) | 0.017 |
| **ED** |  |  |  |  |  |  |
|  | Age | 0.98 (0.95-1.01) | 0.155 | Age | 1.01 (0.99-1.04) | 0.266 |
|  | Male gender | 1.79 (1.04-3.10) | 0.037 | Male gender | 2.98 (1.78-4.98) | <0.001 |
|  | Breathlessness | 0.25 (0.13-0.48) | <0.001 | Brain fog | 0.29 (0.1-0.85) | 0.025 |
|  | Myalgia | 0.26 (0.09-0.75) | 0.013 | Fatigue | 0.40 (0.24-0.67) | 0.001 |
|  | Cough | 2.71 (1.28-5.74) | 0.009 |  |  |  |
|  | Arthralgia | 3.92 (1.12-13.77) | 0.033 |  |  |  |

## Discussion

Our 12-month experience in the earliest post-COVID clinical service in the UK to include NH, PH and PED patients highlights five findings. First, we document significant functional impairment across all patient groups, particularly NH individuals. Second, we identify the need for multidisciplinary, structured assessment following SARS-CoV-2 infection in all patient groups. Third, we show variations in symptoms and diagnostic features across patient groups. Fourth, we describe high burden of specialist input, onward therapy and psychology support in different patient groups in a real-world post-COVID clinical service. Fifth, we identify factors which could contribute to inequitable access to post-COVID care.

Our results underscore the patient and system need for comprehensive measurement and reporting of effects of SARS-CoV-2 infection in all individuals (PH and NH) on health, function, and healthcare utilisation, and the profound ramifications for individuals, their families and communities beyond measures of functional impairment and organ dysfunction. Patients with Long COVID have called for “recognition, research and rehabilitation”(22), but COVID-19 research and care have focused primarily on acute physiology, management, and mortality in hospitalised individuals. It has been previously established that burden of specific diseases can only be compared across population groups when morbidity in addition to mortality is accurately recorded(23). Long COVID burden must be documented in this way.

The extent of multi-morbidity and functional impairment in Long COVID, requires cross-speciality, multi-professional, integrated working, and development of broader clinical expertise, with transferable benefits to other conditions beyond the pandemic(21) rather than siloed, organ-based approaches(19,24). Such integrated care pathways have been effective in other diseases pre-pandemic(25), and could support rapid up-skilling and skills transference needed for the wider workforce in post-COVID clinical care. To enable a consistent, holistic approach, we developed structured EHR tools to capture functional and psychological impact. We implemented multi-speciality, multi-disciplinary meetings to enhance efficiency and share learning, seeking patient feedback from all encounters to improve services. Further analysis of correlation between symptoms, functional consequences and investigations via data-driven approaches are essential to guide mechanistic studies, treatment approaches and risk stratification. Hospital- and clinic-based analyses are important, given slow uptake of new Long COVID coding in primary care(26).

As in other studies, we document severe functional impairment(11-13) and significant mortality risk post-hospital discharge. We also document severe functional impairment in NH and PED individuals despite low predicted(2) and observed mortality risk. UK guidance for post-hospital follow-up prioritised those requiring greater levels of organ support or with more severe chest imaging abnormality(20). This focus risks lack of recognition of morbidity in patients with less severe acute respiratory presentations of SARS-CoV-2 infection. The multivariable models suggested that fatigue, brain fog, chest pain and breathlessness were the common factors for continued suboptimal health and not returning to work. Other factors were less consistently included in the models and some were associated in an unexpected direction, which may be due to a real effect or confounding with other symptoms. Underlying mechanisms and management require further research.

Like other studies, we show prolonged variation in symptoms and extent of diagnostic abnormality between different cohorts, suggesting different phenotypes of post-COVID syndrome/Long COVID(27). Absence of a specific diagnostic test or biomarker(28) makes the balance between adequate investigation of significant symptoms versus “over-medicalising” challenging. For example, elevated D-dimer levels triggered frequent CTPA requests, but with low diagnostic yield in NH patients. Exercise testing showed oxygen desaturation most frequently in PH patients, but also in NH groups without clear evidence of lung or heart abnormalities. Echocardiography rarely showed abnormal findings, despite frequent use to investigate chest pain and breathlessness. The clinical significance of detectable abnormalities on cMRI in patients with chest pain requires further evaluation(29). Future research must determine whether and how diagnostic abnormalities relate to end-organ effects or mechanisms underlying Long COVID.

Patients often needed further specialist opinion and onward referral to community rehabilitation and psychological support, with significant resource implication given such pathways are currently poorly defined. In addition to the ongoing waves of COVID-19 infection, threats of new variants, and effects on non-COVID services(2); the “long tail” of the pandemic in terms of Long COVID is a concern for policy and health service planning in all countries. Rehabilitation needs are complex and likely to require novel multi-modal therapy approaches which integrate psychological support, and workforce capacity and capability which are not currently in place. Expertise in fatigue-management, treatment for disordered breathing patterns and programs to support return to employment are particular priorities. Novel digital self-management solutions could be a useful adjunct but need further evaluation. Although different post–COVID care models are evolving(30,31), the evidence base is minimal with few, large-scale, pragmatic trials of treatment and rehabilitation. Developing integrated, research-oriented pathways in EHR-enabled health systems could provide a platform for rapid evaluation of investigation, treatment and rehabilitation approaches alongside delivering care, which has been effective in acute COVID-19(32).

There are signs of inequitable access to care in our cohort. Although observed ethnicity of NH patients reflected the catchment population, it contrasted with disproportionate burden of acute SARS-CoV-2 in ethnic minorities. NH patients were also less likely to live in areas of deprivation and may have had easier access to referral. Varying referral rates by primary care provider were seen, requiring further investigation to understand contributing factors. Under-diagnosis of Long COVID has been described in primary care in England(31). NH patients had delayed referral. Consideration should be given to proactive follow-up of patients managed in the community, particularly given the severity of illness in patients referred. Follow-up strategies should also account for needs of frail, elderly, co-morbid, and hard-to-reach patients, who were under-represented in our cohort.

Our findings have several limitations. We report outcomes from a single centre, unlikely to be representative of the whole UK population. As post-COVID clinics are established around the UK and in other countries, prospective data collection and comparison will enable variations to be investigated. We used subjective symptom and functional status data, which may have self-reporting bias. We report real-world, clinical findings, therefore data regarding pre-morbid status, baseline characteristics, investigations and follow-up (particularly for patients with persistent disease for 12 months or more) are more limited than in dedicated research cohorts. NH patients were self-selecting whilst PH patients were included as part of routine follow-up. This selection bias precludes both making precise estimates of the burden of Long COVID amongst NH patients and also making comparisons between the NH patients and other patient groups. In our basic logistic regression models, we modelled age as a linear variable, which may have obscured true non-linear effects of age in these models. Our models were based on cross-sectional data at presentation to the clinic.

Our findings have several implications for research and policy. First, we identify differences in longer-term effects of SARS-CoV-2 infection in PH versus individuals, supporting the existence of a number of different phenotypes of Long COVID. Further correlation of symptom clusters, functional impacts and diagnostic information is required to better define phenotypes and to inform studies of underlying mechanisms and potential treatments. Second, a wide range of patient-reported outcome measures were required to capture the impact of Long COVID on individuals. Development of a validated clinical assessment tool will further enable investigation of the natural history of the condition across larger data-sets and differing EHR systems. Third, we identify a need for a novel model of rehabilitation, incorporating psychological support and urgent solutions to address the current workforce shortfalls. Fourth, the multi-system nature of the condition will require broadening of clinical expertise both within primary and secondary care and effective integrated pathways. Finally, given the severity of illness documented in all patient groups, our analysis supports the need for swift, equitable, access to assessment for all patients suffering ongoing illness after SARS-CoV-2 infection, as well as effective infection suppression policy for all.

Post-COVID morbidity can be severe, regardless of severity of acute illness, and scale of healthcare utilisation and inability to return to employment represent a major burden to individuals, healthcare and welfare systems, and economies. Definition of Long-COVID phenotypes and development and evaluation of diagnostic, treatment and rehabilitation approaches is urgently required. Dissemination of clinical expertise in management of Post-COVID complications needs to occur across integrated care systems and policy change is required to improve equity of access by individuals to appropriate levels of care and support.

## Data Availability

All data relevant to the report is included in this manuscript and supplementary material.

## Contributors

MH has led the UCLH Post-COVID service since inception, with TH. MM, RL, HR, RB, MZ, PN, EW, RE, AC, EB, PM, ED, provided clinical support to the service. MH, TH and AB wrote the first and final drafts of the manuscript. JP conducted analyses, with input from MH, SC and AB, and statistical input from HM-D and GP. All authors provided critical feedback on earlier and final drafts of the manuscript.

## Data sharing

All data relevant to the report is included in this manuscript and supplementary material.

## Acknowledgements

N/A

