## Supplementary methods, tables and figures for "Post-COVID assessment in a specialist clinical service: a 12-month, single-centre analysis of symptoms and healthcare needs in 1325 individuals"

**Table of Contents**

|  | **Page** |
| --- | --- |
| **Methods Supplement.** | **3,4** |
| **Web Figure 1.** Index of multiple deprivation deciles in 1321 individuals referred to the post-COVID assessment clinic by referral source. | **5** |
| **Web Figure 2.** Referral rate for post- COVID assessment and size of general practice population. | **6** |
| **Web Figure 3.** Reported percentage of normal health in individuals referred to the post-COVID assessment clinic at first assessment. | **7** |
| **Web Figure 4.** Fatigue Assessment Scale score in 806 individuals referred to the post-COVID assessment clinic at first assessment. | **8** |
| **Web Figure 5.** Frequency of symptoms at first assessment in 1325 individuals referred to the post-COVID assessment clinic. | **9** |
| **Web Table 1.** Work patterns in 1325 individuals referred to the post-COVID assessment clinic at at first assessment. | **10** |
| **Web Table 2.** Blood investigations at first assessment in individuals seen in the post-COVID assessment clinic. | **11** |
| **Web Table 3.** Visual Analogue Scale scores at first assessment in individuals referred to the post-COVID assessment clinic. | **12** |

**Methods Supplement**

**Index of Multiple Deprivation (IMD) decile**

We used IMD decile as a marker of relative deprivation, determined according to 2019 data from the United Kingdom Ministry of Housing, Communities and Local Government ^[1]^. This provides data based on a post-code that typically contains around 15 households.

**Fatigue Assessment Scale (FAS)**

Patients were assessed, according to clinical judgement, using the Fatigue Assessment Scale (FAS) (n=806)^[2]^.

**Blood investigations**

Patients were subject to blood investigations at presentation according to clinical judgement (Alanine transaminase (ALT) **n=615**, Creatine kinase (CK) **n= 521**, Cardiac Troponin T **n=558**, Full blood count **n=601**, D-Dimer **n=567**, NT-proBNP **n=239**). Local reference ranges were used to determine raised values:

|  | **Upper limit of normal** |
| --- | --- |
| **ALT** (U/L) | 35 (Females); 50 (Males) |
| **CK** (U/L) | 140 (Females); 204 (Males) |
| **Cardiac Troponin T** (ng/L) | 14 |
| **Eosinophils** (x 10^9^/L) | 0.40 |
| **D-Dimer μg/L FEU** | 550 |
| **NT-proBNP ng/L** | 400 |

Results for CK and ALT are stratified by gender due to differing reference ranges.

**Return to work status**

Patients were asked ‘Would you feel able to return to work if permitted?' during the consultation. Possible responses were ‘Full time’, ‘Part time’, ‘Not at all’, ‘Retired’ or ‘Not employed’. Where this information was not available, the response was recorded as ‘Not stated’. Percentages are reported as a proportion of all employed individuals for individuals who were employed, and as a proportion of all individuals where employment status was unemployed, retired or unknown.

**One Minute Sit to Stand Exercise Capacity Test**

The test can be conducted in a standard consultation room, and requires only a saturation probe and a standard office chair as equipment.

The chair seat height should be 46cm.

During a sit to stand repetition, the patient's bottom should touch the seat of the chair, but the full weight of the patient does not have to be placed on the chair.

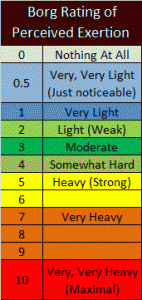

**Sit to stand protocol:**

- A saturation probe is placed on the patient's finger
- The resting heart rate and saturations are recorded
- A modified Borg rating is taken (see chart opposite)
- The patient is asked to perform as many sit to stand repetitions as they are able in 60 seconds
- The saturations and heart rate are monitored throughout.
- The test is stopped if the oxygen saturations drop by ≥4% from baseline
- The test is stopped if the patient reports discomfort or is unable to continue
- At the point of stopping the heart rate, saturations and Borg score are taken again.
- Record the number of repetitions achieved.
- The time to recovery, minimum saturations, and maximum heart rate are noted.

**Web figure 1.** Index of multiple deprivation deciles in 1321 individuals referred to the post-COVID assessment clinic by referral source.

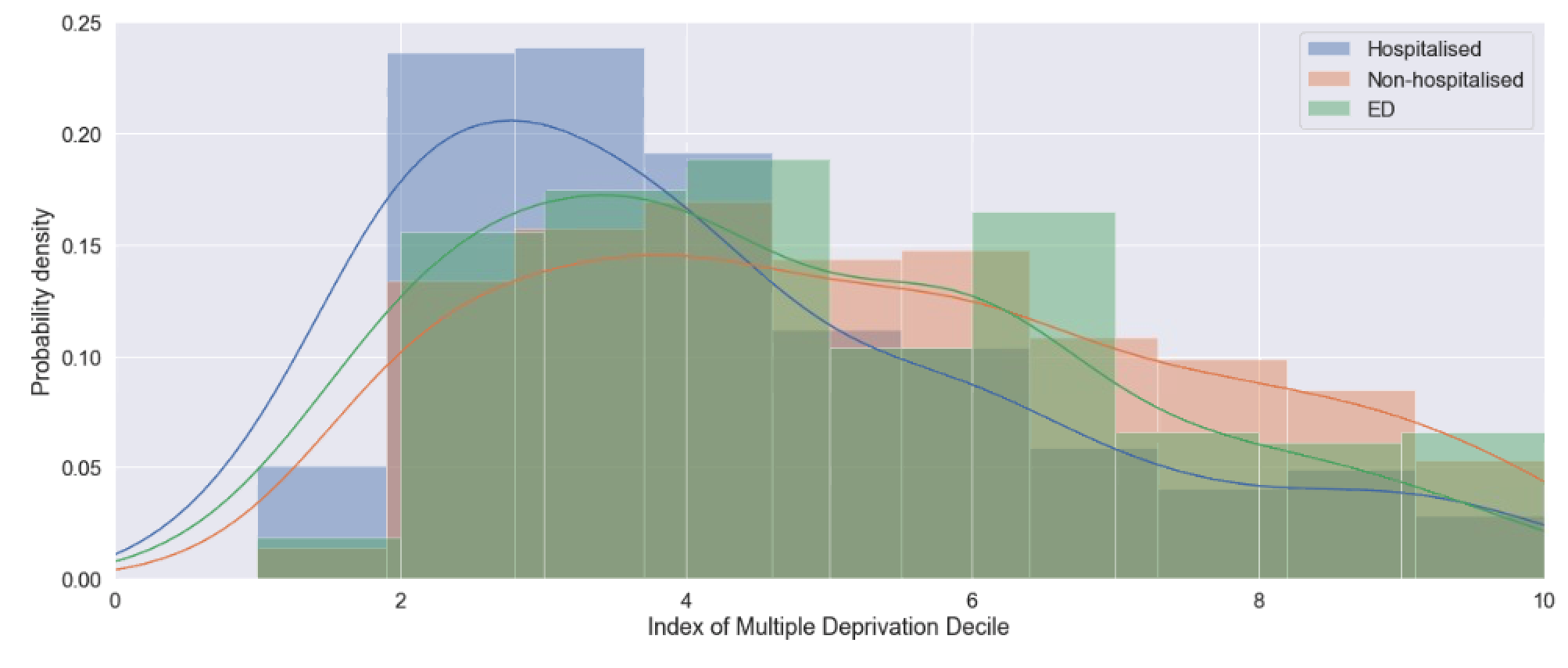

**Web Figure 2.** Referral rate for post-COVID assessment and size of general practice population.

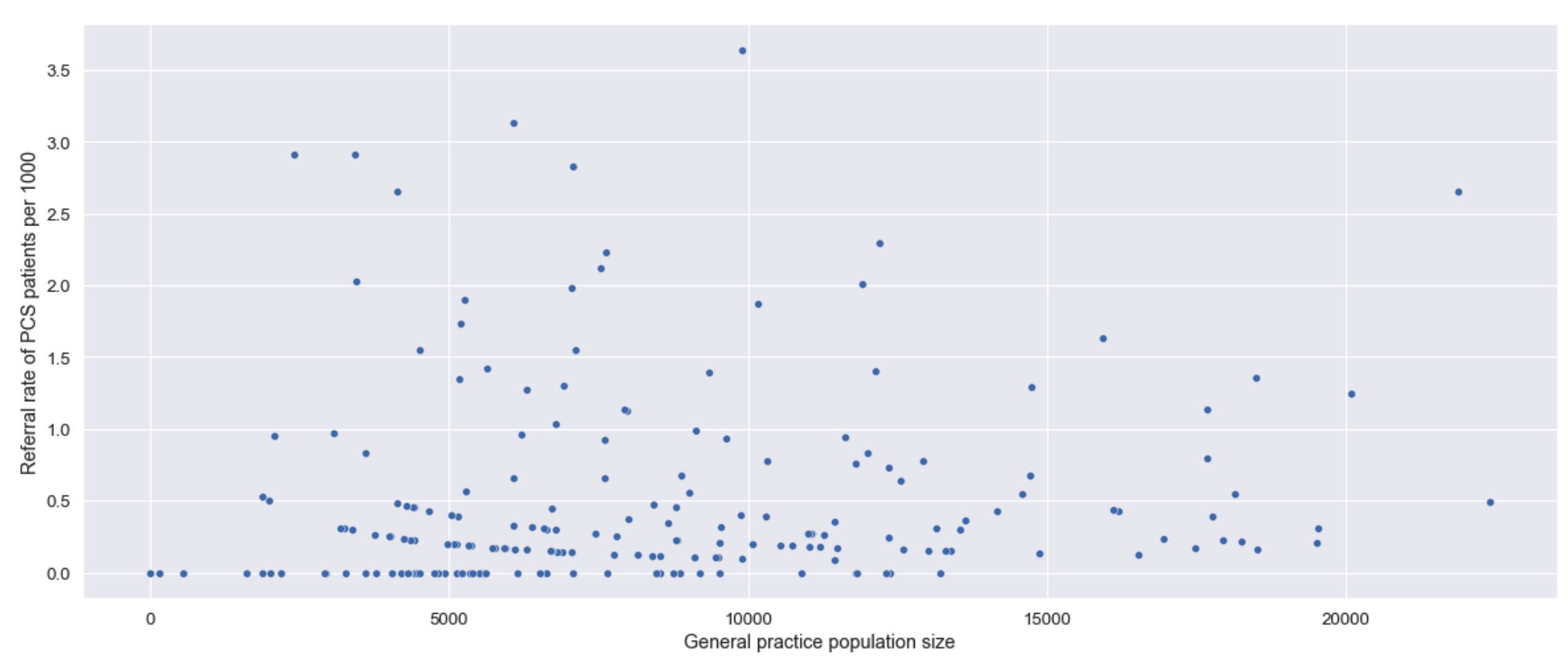

**Web Figure 3.** Reported percentage of normal health in individuals referred to the post-COVID assessment clinic at first assessment.

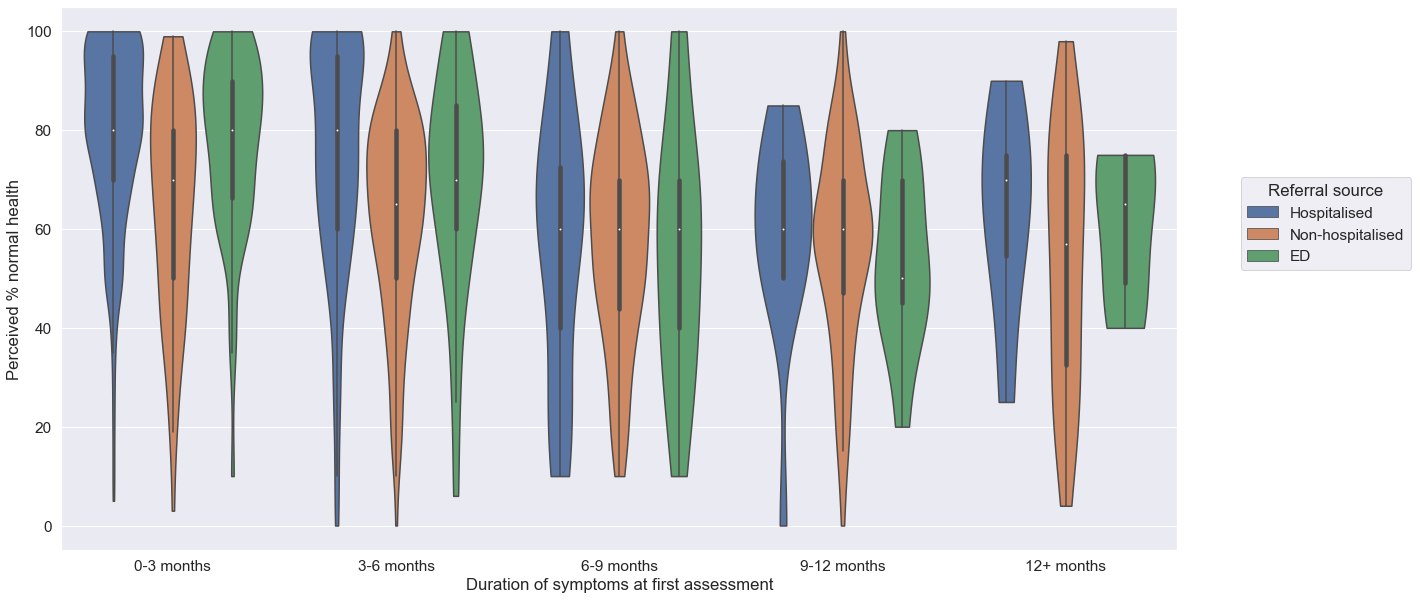

**Web Figure 4.** Fatigue Assessment Scale score in 806 individuals referred to the post-COVID assessment clinic at first assessment.

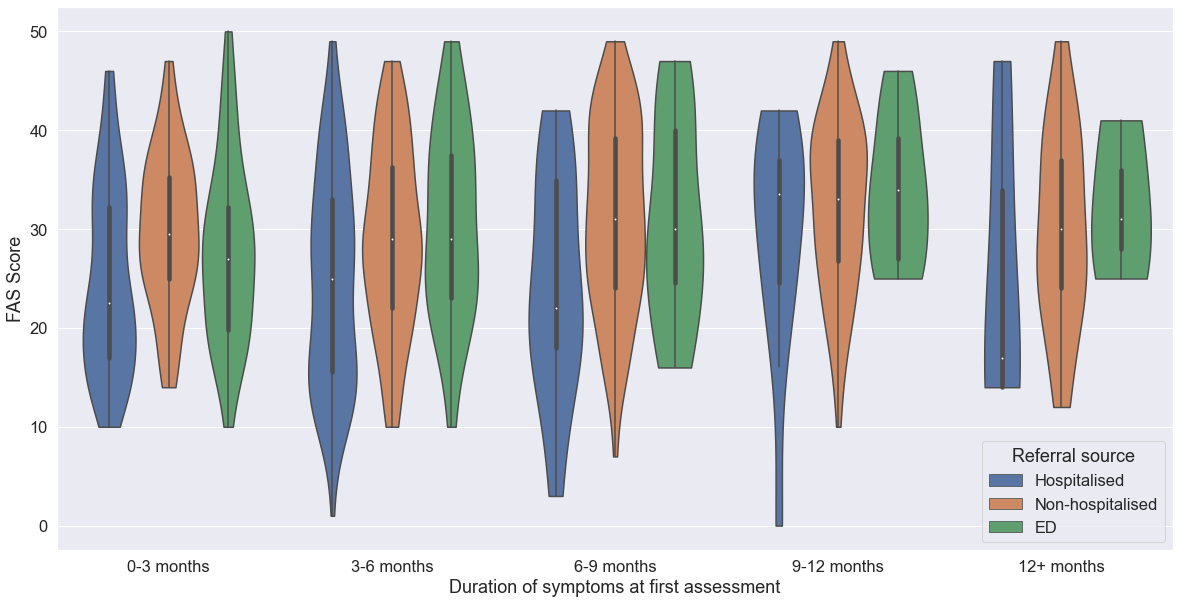

**Web Figure 5.** Frequency of symptoms at first assessment in 1325 individuals referred to the post-COVID assessment clinic.

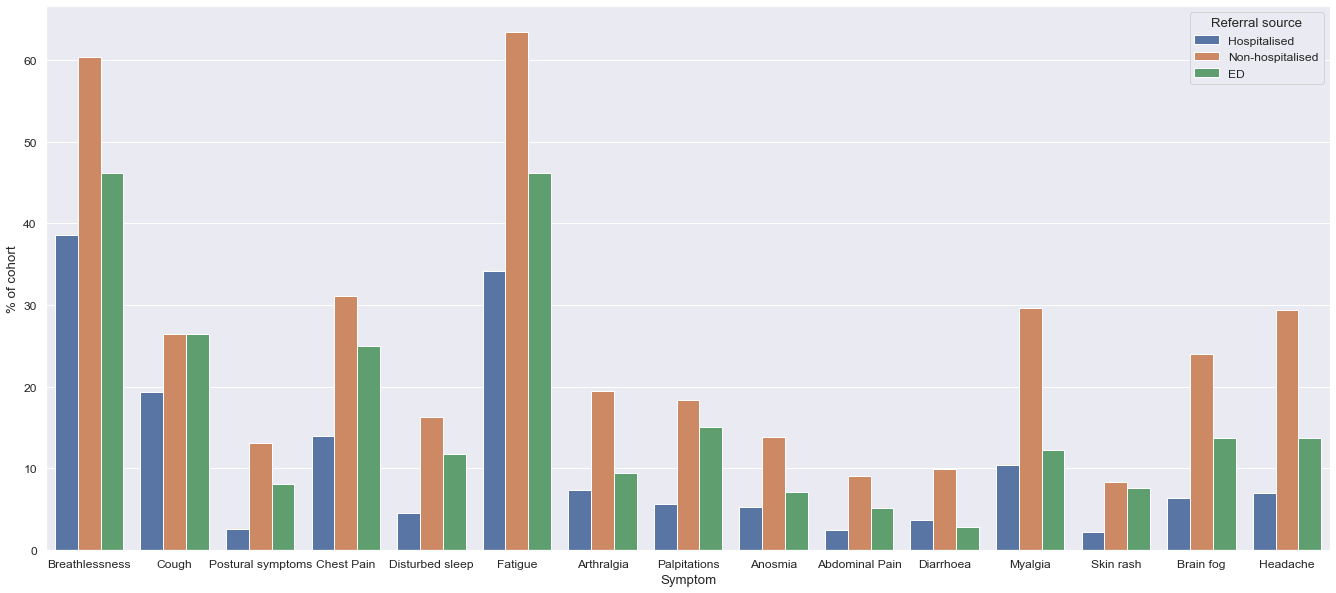

**Web Table 1:** Work patterns in 1325 individuals referred to the post-COVID assessment clinic at first assessment.

**Hospitalised (n=547)**

|  | **0-3 months (n=356)** | **3-6 months (n=151)** | **6-9 months (n=19)** | **9-12 months (n=14)** | **12+ months (n=7)** |
| --- | --- | --- | --- | --- | --- |
| **Employed** | **215 (60.4%)** | **95 (62.9%)** | **16 (84.2%)** | **13 (92.9%)** | **5 (71.4%)** |
| *Full time* | *93 (43.3%)* | *42 (44.2%)* | *6 (37.5%)* | *4 (30.8%)* | *2 (40.0%)* |
| *Part time* | *54 (25.1%)* | *19 (20.0%)* | *3 (18.8%)* | *1 (7.7%)* | *2 (40.0%)* |
| *Not at all* | *68 (31.6%)* | *34 (35.8%)* | *7 (43.8%)* | *8 (61.5%)* | *1 (20.0%)* |
| **Not employed** | **44 (12.4%)** | **15 (9.9%)** | **2 (10.5%)** | **(0.0%)** | **1 (14.3%)** |
| **Retired** | **97 (27.2%)** | **41 (27.2%)** | **1 (5.3%)** | **1 (7.1%)** | **1 (14.3%)** |
| **Not stated** | **0 (0.0%)** | **0 (0.0%)** | **0 (0.0%)** | **0 (0.0%)** | **0 (0.0%)** |

**Non-hospitalised (n=566)**

|  | **0-3 months (n=77)** | **3-6 months (n=183)** | **6-9 months (n=128)** | **9-12 months (n=143)** | **12+ months (n=35)** |
| --- | --- | --- | --- | --- | --- |
| **Employed** | **67 (87.0%)** | **167 (91.3%)** | **118 (92.3%)** | **124 (86.7%)** | **30 (85.7%)** |
| *Full time* | *23 (34.3%)* | *70 (41.9%)* | *54 (45.8%)* | *49 (39.5%)* | *13 (43.3%)* |
| *Part time* | *20 (29.9%)* | *56 (33.5%)* | *34 (28.8%)* | *37 (29.8%)* | *9 (30.0%)* |
| *Not at all* | *24 (35.8%)* | *41 (24.6%)* | *30 (25.4%)* | *38 (30.6%)* | *8 (26.7%)* |
| **Not employed** | **7 (9.1%)** | **11 (6.0%)** | **6 (4.7%)** | **14 (9.8%)** | **2 (5.7%)** |
| **Retired** | **3 (3.9%)** | **5 (2.7%)** | **4 (3.1%)** | **5 (3.5%)** | **3 (8.6%)** |
| **Not stated** | **0 (0.0%)** | **0 (0.0%)** | **0 (0.0%)** | **0 (0.0%)** | **0 (0.0%)** |

**ED (n=212)**

|  | **0-3 months (n=126)** | **3-6 months (n=53)** | **6-9 months (n=17)** | **9-12 months (n=11)** | **12+ months (n=5)** |
| --- | --- | --- | --- | --- | --- |
| **Employed** | **107 (84.9%)** | **46 (86.8%)** | **16 (94.1%)** | **6 (54.5%)** | **3 (60.0%)** |
| *Full time* | *50 (46.7%)* | *19 (41.3%)* | *7 (43.8%)* | *3 (50.0%)* | *3 (100.0%)* |
| *Part time* | *30 (28.0%)* | *17 (37.0%)* | *5 (31.2%)* | *(0.0%)* | *(0.0%)* |
| *Not at all* | *27 (25.2%)* | *10 (21.7%)* | *4 (25.0%)* | *3 (50.0%)* | *(0.0%)* |
| **Not employed** | **10 (7.9%)** | **3 (5.7%)** | **(0.0%)** | **5 (45.5%)** | **1 (20.0%)** |
| **Retired** | **9 (7.1%)** | **4 (7.5%)** | **1 (5.9%)** | **(0.0%)** | **1 (20.0%)** |
| **Not stated** | **0 (0.0%)** | **0 (0.0%)** | **0 (0.0%)** | **0 (0.0%)** | **0 (0.0%)** |

* Percentages are reported as a proportion of all employed individuals for individuals who were employed, and as a proportion of all individuals where employment status was unemployed, retired or unknown.

**Web Table 2:** Blood investigations at first assessment in individuals referred to the post-COVID assessment clinic.

|  | **Overall (N=1325)** | **Hospitalised (n=547)** | **Non-hospitalised (n=566)** | **ED (n=212)** | **p** |
| --- | --- | --- | --- | --- | --- |
| **ALT (Males) Normal Range 10-50 IU/L** |  |  |  |  |  |
| **n** | **316 (23.8%)** | **162 (29.6%)** | **99 (17.5%)** | **55 (25.9%)** |  |
| Median (IQR) | 31 (22-45) | 30 (21-45.75) | 32 (24-44.5) | 31 (24-45) | 0.6 |
| N (%) raised | 60 (19.0%) | 33 (20.4%) | 16 (16.2%) | 11 (20.0%) | |
| **ALT (Females) Normal range 10-35 IU/L** |  |  |  |  |  |
| **n** | **299 (22.6%)** | **94 (17.2%)** | **144 (25.4%)** | **61 (28.8%)** |  |
| Median (IQR) | 21 (17-29) | 21 (18-28.5) | 21 (17-27.5) | 23 (17-29) | 0.47 |
| N (%) raised | 51 (17.1%) | 21 (22.3%) | 24 (16.7%) | 6 (9.8%) |  |
| **CK (Males) Normal range 38-204 IU/L** |  |  |  |  |  |
| **n** | **278 (21.0%)** | **139 (25.4%)** | **86 (15.2%)** | **53 (25.0%)** |  |
| Median (IQR) | 119 (81-180.75) | 102 (69-163) | 124.5 (90.75-172.25) | 147 (99-208) | **< 0.001** |
| N (%) raised | 54 (19.4%) | 22 (15.8%) | 17 (19.8%) | 15 (28.3%) | |
| **CK (Females) Normal range 26-140 IU/L** |  |  |  |  |  |
| **n** | **243 (18.3%)** | **74 (13.5%)** | **119 (21.0%)** | **50 (23.6%)** |  |
| Median (IQR) | 82 (63-114.5) | 77 (50.75-104) | 82 (69-120) | 86.5 (70.25-116.25) | 0.07 |
| N (%) raised | 34 (14.0%) | 7 (9.5%) | 20 (16.8%) | 7 (14.0%) |  |
| **Eosinophils Normal range 0.0 – 0.4 x10^9^/L** |  |  |  |  |  |
| **n** | **601 (45.4%)** | **249 (45.5%)** | **239 (42.2%)** | **113 (53.3%)** |  |
| Median (IQR) | 0.13 (0.08-0.21) | 0.15 (0.10-0.23) | 0.11 (0.06-0.18) | 0.12 (0.08-0.26) | **< 0.001** |
| N (%) raised | 35 (5.8%) | 17 (6.8%) | 6 (2.5%) | 12 (10.6%) | |
| **D-Dimer Normal range 0-550 μg/L FEU** |  |  |  |  |  |
| **n** | **567 (42.8%)** | **230 (42.0%)** | **234 (41.3%)** | **103 (48.6%)** |  |
| Median (IQR) | 290 (210-470) | 365 (250-587.5) | 250 (190-340) | 320 (225-470) | **< 0.001** |
| N (%) raised | 107 (18.9%) | 65 (28.3%) | 24 (10.3%) | 18 (17.5%) | |
| **NT-proBNP Normal range 0-400 ng/L** |  |  |  |  |  |
| **n** | **239 (18.0%)** | **112 (20.5%)** | **93 (16.4%)** | **34 (16.0%)** |  |
| Median (IQR) | 93 (66-164) | 120 (74.25-256.25) | 86 (62-109) | 79 (63.25-106.75) | **< 0.001** |
| N (%) raised | 19 (7.9%) | 19 (17.0%) | 0 (0.0%) | 0 (0.0%) |  |
| **Cardiac Troponin T Normal Range 0-14 ng/L** | |  |  |  |  |
| **n** | **558 (42.1%)** | **215 (39.3%)** | **234 (41.3%)** | **109 (51.4%)** |  |
| Median (IQR) | 6 (4-9) | 8 (5-14) | 5 (4-7) | 6 (4-7) | **< 0.001** |
| N (%) raised | 75 (13.4%) | 58 (27.0%) | 11 (4.7%) | 6 (5.5%) |  |

**Web Table 3.** Visual Analogue Scale scores at first assessment in individuals referred to the post-COVID assessment clinic.

|  | **Breathlessness** | | **Fatigue** | | **Cough** | | **Sleep quality** | | **Palpitations** | |
| --- | --- | --- | --- | --- | --- | --- | --- | --- | --- | --- |
|  | *n (%)* | *Median (IQR)* | *n (%)* | *Median (IQR)* | *n (%)* | *Median (IQR)* | *n (%)* | *Median (IQR)* | *n (%)* | *Median (IQR)* |
| Overall (n=1325) | 893 (67.4%) | 4 (1-6) | 894 (67.5%) | 5 (3-7) | 870 (65.7%) | 0 (0-2) | 850 (64.2%) | 4 (1-7) | 772 (58.3%) | 0 (0-4) |
| Hospitalised (n=547) | 308 (56.3%) | 3 (0-5) | 304 (55.6%) | 4 (1-6) | 298 (54.5%) | 0 (0-2) | 285 (52.1%) | 2 (0-6) | 248 (45.3%) | 0 (0-1) |
| Non-hospitalised (n=566) | 474 (83.7%) | 4 (2-6) | 478 (84.5%) | 6 (4-8) | 464 (82.0%) | 0 (0-3) | 460 (81.3%) | 5 (2-7) | 430 (76.0%) | 2 (0-5) |
| ED (n=212) | 111 (52.4%) | 3 (1-6) | 112 (52.8%) | 5 (2-7) | 108 (50.9%) | 0.5 (0-3) | 105 (49.5%) | 4 (1-7) | 94 (44.3%) | 2 (0-4.75) |

1. English indices of deprivation 2019: Postcode Lookup [Internet]. Imd-by-postcode.opendatacommunities.org. 2021 [cited 22 May 2021]. Available from: https://imd-by-postcode.opendatacommunities.org/imd/2019
2. Michielsen H, De Vries J, Van Heck G. Psychometric qualities of a brief self-rated fatigue measure. Journal of Psychosomatic Research. 2003;54(4):345-352.
